# A mixed-methods assessment of malaria case investigations and response in the elimination setting of Southern Province, Zambia

**DOI:** 10.64898/2026.05.23.26353921

**Authors:** Refilwe Y. Karabo, Salima M. Kalyalya, John Miller, Kafula Silumbe, Busiku Hamainza, Chris Lungu, Javan Chanda, Arantxa Roca-Feltrer, Adam Bennett, Caterina Guinovart, Zhiyuan Mao, Ruth A. Ashton, Jeni A. Stolow, Thomas P. Eisele

## Abstract

**Background:** In 2017, Zambia adopted surveillance as a core intervention towards achieving malaria elimination. Among the surveillance strategies is the malaria case investigation and response 1-3-7 (MCIR 1-3-7), which has been piloted in two low-incidence districts in the Southern Province since 2021. The study aimed to assess the implementation of MCIR 1-3-7 under programmatic conditions. It examined the timeliness, and completeness of the MCIR 1-3-7 activities, including the completeness of data entry in surveillance forms, and explored the experiences and perspectives of healthcare workers involved in the pilot.

**Methods:** A mixed-methods design was employed to assess the MCIR 1-3-7. Using a descriptive cross-sectional design, quantitative data were collected from 19 healthcare facilities in the two districts to assess the timeliness and completeness of MCIR 1-3-7. Additionally, 12 qualitative interviews were conducted with 29 healthcare workers from 11 of the 19 healthcare facilities. The interviews were voice-recorded and then transcribed manually. A codebook was developed using an iterative process to explore the facilitators and barriers encountered by healthcare workers in implementing the MCIR 1-3-7 intervention. All the visited facilities were purposively selected based on logistical convenience.

**Results:** This study retrospectively assessed 510 malaria cases that were diagnosed between January 2022 and June 2023, presenting at 19 health facilities: 283 cases in Chikankata and 227 in Mazabuka districts. A total of 278 cases (54.5%) were deemed to have been imported from outside the district, province, or country, while 45.5% (232/510) of the cases were classified as transmitted locally. Overall, 29.6% of case notification forms were found to be complete. Twelve interviews with 29 healthcare workers revealed a lack of transportation modalities as the main obstacle in executing the MCIR 1-3-7 intervention. The healthcare workers also indicated that monetary incentives, and supportive supervision would help them succeed in implementing this intervention.

**Conclusions:** The MCIR 1-3-7 has the potential to accelerate elimination in areas with low-transmission of malaria in Zambia. This study highlights opportunities to improve future implementation of the MCIR 1-3-7 intervention via strengthening supportive supervision, availing job aids, and ensuring access to malaria commodities as the intervention expands.

## Background

Zambia established the goal of eliminating malaria in 2017 with the launch of the National Malaria Elimination Strategic Plan (NMESP) 2017-21 and the rebranding of the National Malaria Control Center (NMCC) to the National Malaria Elimination Center (NMEC) (1). This reposition was informed by, among others, the significant progress in reducing malaria incidence to very low levels in some districts, increased domestic funding towards curbing malaria, renewed political and financial commitments within the Southern African Development Community (SADC), and the SADC Elimination 8 bloc’s coordinated efforts in cross-border monitoring and control of malaria, as well as cumulative evidence on approaches to addressing malaria and newly available elimination strategies (2).

The Zambian government launched the multi-sectoral End Malaria Council (EMC) in April 2018 to serve as a high-level advocacy plaeorm for malaria elimination, mobilize finances to support elimination efforts, and promote accountability for the implementation of the national strategic plan across all stakeholders (3).

The Ministry of Health subsequently adopted Pillar 3 of the WHO Global Technical Strategy (GTS) for Malaria 2016-2030 in the NMESP 2017-2021 (2). The GTS Pillar 3 aims to “transform malaria surveillance into a core intervention for elimination” (4). In Zambia, this pillar was applied to areas with low malaria transmission, namely Levels 0 and 1 of the malaria epidemiological levels 0-4, where Level 0 represents areas with no malaria, and level 1 is indicated by 1-49 cases per 1,000 population, with no detectable parasite prevalence (5). Adoption of this pillar is further evident in the NMESP 2022-26 (6). Many countries, including Tanzania and South Africa, have adopted the same GTS pillar to help achieve their ambitions for malaria elimination (4),(7),(8),(9).

In 2021, to commit to the malaria elimination efforts, the NMEC, PATH, MACEPA, and other partners embarked on a pilot project of Malaria Case Investigation and Response 1-3-7 (MCIR 1-3-7). The pilot was part of an intervention package to strengthen surveillance in low malaria transmission areas. The MCIR 1-3-7 encompasses the activities of Component D of the Elimination Agenda 2017 (Components A, B, C, D, and E) (10), which involve detecting and investigating individual cases through case investigations in households and villages to stop malaria transmission in areas under epidemiological Level 1. The pilot was conducted in Mazabuka and Chikankata districts of Southern Province, where there is a low malaria prevalence of 3.3% among under-five-year-olds as determined by the rapid diagnostic test in the 2021 Malaria Indicator Survey (11). The two districts are described as approaching elimination.

China was the first country to implement the 1-3-7 approach (12) and attributes its success in eliminating malaria to this strategy, alongside other coordinated efforts (13). Other countries have also adopted this surveillance strategy. According to *Yi and colleagues* (2023) (14), Thailand, Cambodia, and the Lao People’s Democratic Republic have implemented the 1-3-7 approach as part of their surveillance efforts. In sub-Saharan Africa, Tanzania championed this approach, with the assistance of the China-Africa Health Cooperation, and implemented it in the southern part of the country (15). Senegal has also invested in surveillance and adopted the 1-3-7 approach, implementing it in the northern region (16).

In 2022, after one year of implementation in the two pilot districts of Southern Province, the NMEC and stakeholders sought to assess its implementation under routine programmatic conditions. In addition, they sought to garner lessons learned from the pilot districts to guide the NMEC’s planned adoption and scaling up of the MCIR 1-3-7 approach in other low-incidence districts in Zambia.

This study aimed to assess the quantitative and qualitative aspects of the MCIR 1-3-7 intervention, in order to identify gaps, explore potential improvements, and inform decision-making related to its implementation, adoption, and expansion to other low malaria transmission areas. Key outcomes of the quantitative component of this study will help implementing organizations understand whether the intervention was executed on time, whether the 1-3-7 process was done for all eligible cases, and to assess data quality and completeness. The qualitative component of the study will provide insight into the implementers’ perspectives on their ability to execute the activities of the intervention, and how improvements might be achieved.

### MCIR 1-3-7 intervention and procedures

In Zambia, the MCIR 1-3-7 strategy includes timely case notification and reporting within 1 day, household investigations and parasitemia screening, as well as investigations in neighboring households within a 140m radius within 3 days. Lastly, foci investigation and response are conducted within 7 days to curtail further malaria transmission in the foci. A summary of this process is illustrated in Figure 1. MCIR 1-3-7 aims to identify, treat, and track all cases, classify the cases, and ultimately integrate appropriate interventions to halt malaria transmission within the active foci. The case classification is based on reported travel outside the two districts within 30 days prior to the onset of illness and subsequent diagnosis. The main implementers of this intervention are the Community Health Workers (CHWs) who receive support from Environmental Health Technicians (EHTs) and other clinical staff (e.g., nurses) based in the respective healthcare facilities within their catchment areas.

**Fig 1.**
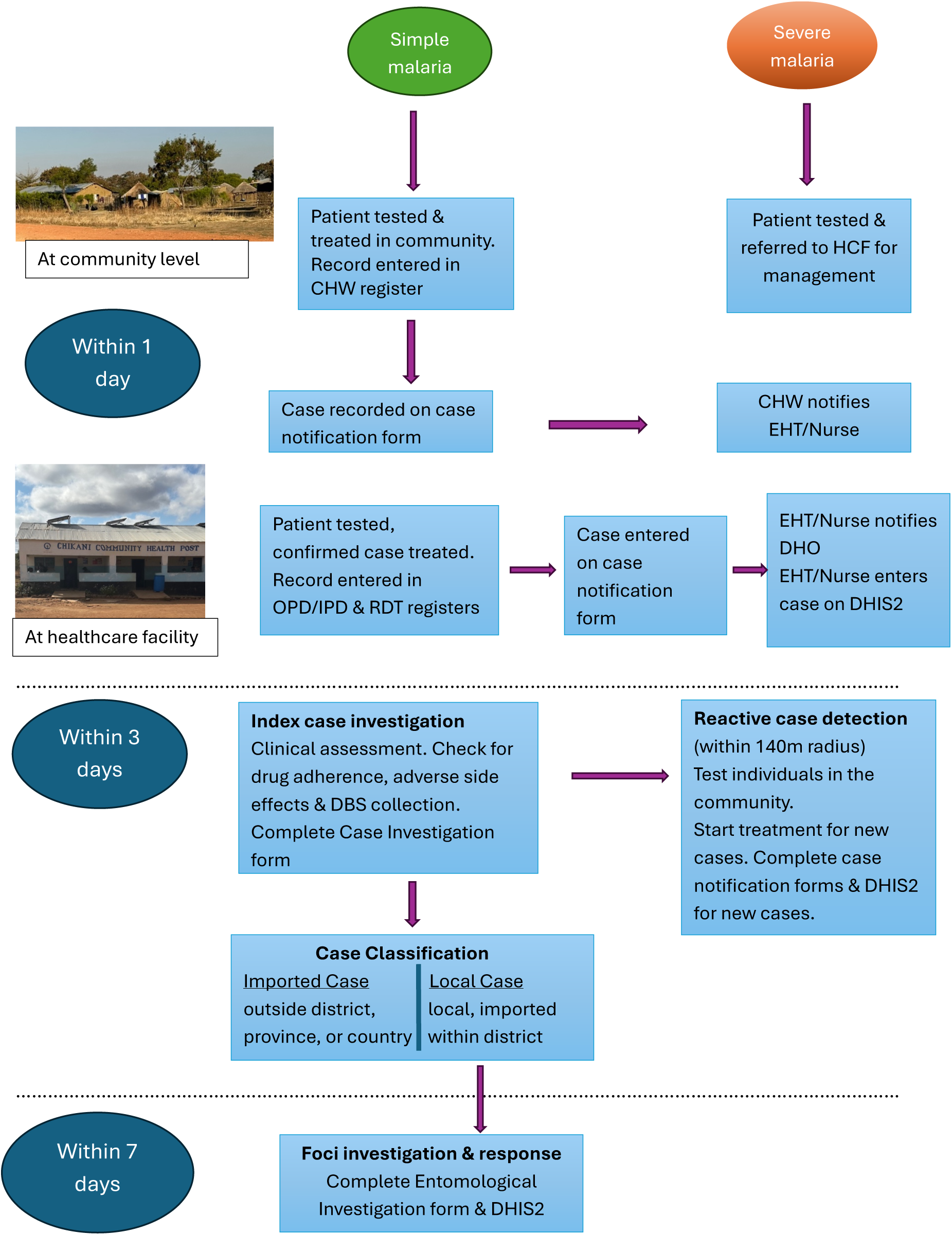
Workflow Implementation of MCIR 1-3-7. *CHW-Community Health Worker, DBS-Dried Blood Spot, DHIS2-District Health Information System, DHO-District Health Office, EHT-Environmental Health Technician, HCF-Healthcare Facility, IPD-In-Patient Department, OPD-Out-Patient Department, RDT-Rapid Diagnostic Test

The healthcare workers record the MCIR 1-3-7 data in four MCIR 1-3-7 paper forms [included in the supplemental file]. The clinical staff also enter the data in the ‘open source, web-based platform’ District Health Information System 2 (DHIS2) (17). Zambia adopted the use of DHIS2 for malaria in 2011 (18), enabling the collection and management of both aggregate and case-based data. The same variables (e.g., demographics, treatment, travel history, and entomological information) that appear on the four MCIR 1-3-7 forms, are available on the DHIS2 digital case-based platform in the selected districts that implement the MCIR 1-3-7.

## Methods

### Study design

Quantitative data were collected between 10-20 July 2023, by a research team from Tulane University. The study utilized a mixed-methods approach, collecting both quantitative and qualitative data simultaneously to acquire a more comprehensive understanding of the intervention. The quantitative component employed a descriptive cross-sectional design to evaluate timeliness and completeness of the surveillance activities. Data were collected from 19 healthcare facilities. The sampled cases had been diagnosed and treated for malaria between January 2022 and June 2023. This period encompassed two peak malaria transmission periods in 2022 and 2023, which run from April to May each year. The qualitative component involved conducting 12 interviews at 11 of the 19 healthcare facilities. The data were collected between 10-17 July 2023. The qualitative and quantitative methods were co-analyzed via a convergent parallel design.

### Selection and eligibility of the facilities for the study

The MCIR 1-3-7 pilot began in October 2021. To be eligible for participation in this study, facilities in Chikankata and Mazabuka must have registered, on average, more than one malaria case per month in 2021 so that a sufficient number of cases could be included. The selected facilities in Mazabuka registered between 12 and 94 confirmed cases, while healthcare facilities in Chikankata had 12-143 confirmed malaria cases each month in 2021. Among eligible facilities, they were also purposively selected based on logistical convenience, including road accessibility and proximity to the researchers’ lodging (Fig 2 and Fig 3).

**Fig 2.**
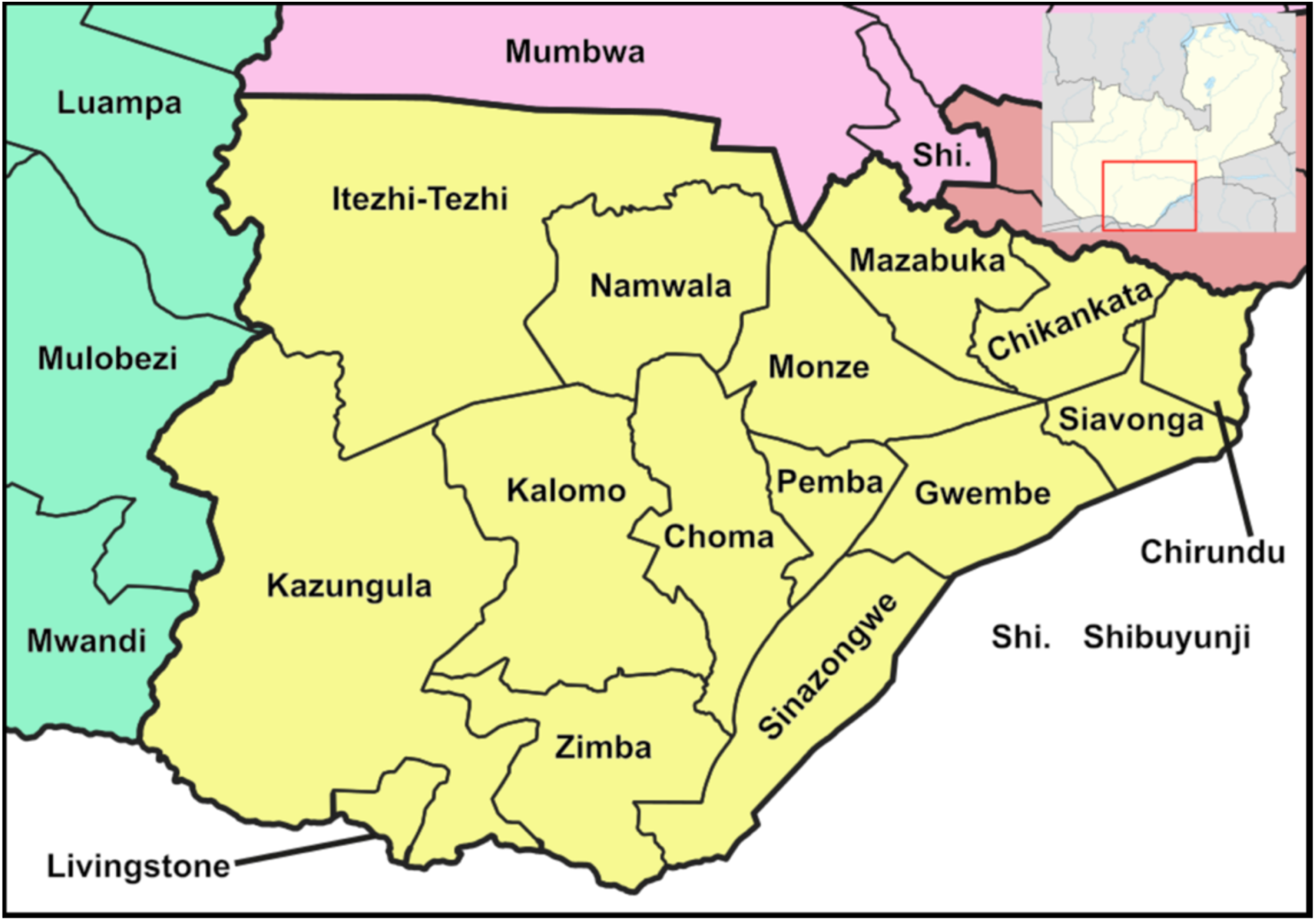
Map of Southern Province, Zambia. Furfur, Peter in s, Location map:NordNordWest - Zambia districts 2022.svg,

**Fig 3.**
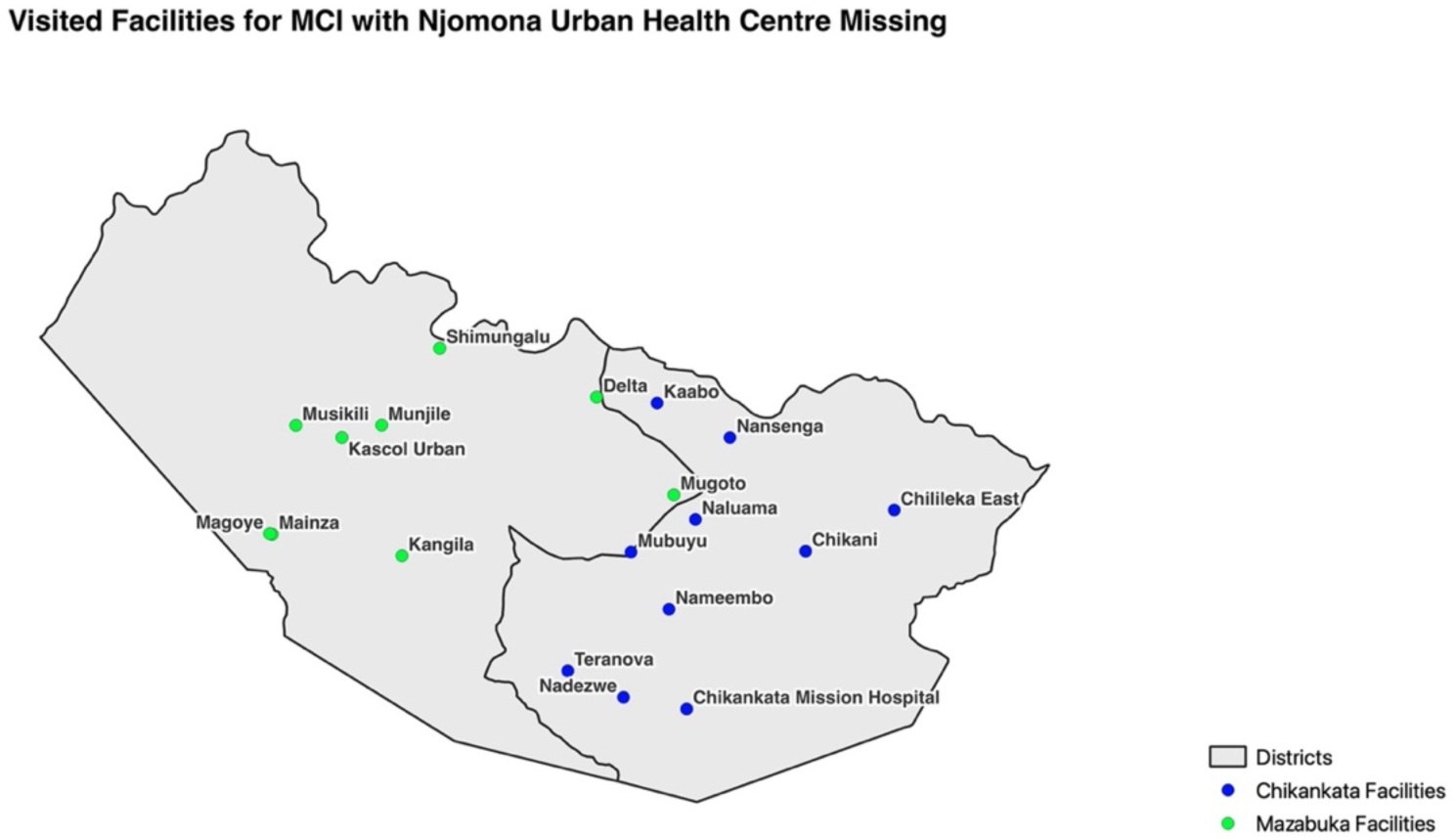
Healthcare facilities visited for data collection.

### Data collection procedures

#### Quantitative data

Data for the quantitative method were collected from paper-based and electronic platforms. From the case notification form, the extracted data included patient name, age, case identification, sex, date of the patient’s visit, diagnosis, and case classification (local/imported). All indicators were examined on paper forms, while DHIS2 indicators were limited to patient name, sex, case identification, case classification, and diagnosis.

From the case investigation form I, the researchers collected data on the follow-up date, the number of people in the household, and the number of people tested for malaria. The case investigation form II provided data on the follow-up date, and the details of a new malaria case. The healthcare workers completed this form only when an individual tested positive for malaria during a visit to the home of an index case. On the entomological investigation form, the researchers collected data on the foci investigation date, the recommended and planned actions for response, and the interventions implemented by day 7. The research team inspected the forms to determine whether they were completed in full.

#### Qualitative data

Twelve interviews were conducted with 29 key informants across the two districts, each lasting approximately one hour. The researchers conducted the interviews in both Nyanja and English to accommodate the language preferences of the interviewees. Audio recordings of all interviews were obtained using a portable voice recorder to capture the exchange between interviewers and interviewees. Verbal consent was obtained from all the interviewees, and permission to audio-record was requested at the beginning of each interview. Participants were informed that their participation was voluntary and that they could decline to respond to any question they felt uncomfortable answering. They were also assured that their identities would remain confidential and would not be linked to their responses or disclosed to the NMEC. Data saturation was reached when the interviews no longer provided new information, as determined by the research team’s consensus.

#### Ethical Approval

Ethical approval was obtained from the University of Zambia Biomedical Research Ethics Committee (UNZABREC). The PATH Research Committee determined that this assessment did not meet the Human Research Protection and IRB application processes, and duly informed Tulane University accordingly. Written consent was not obtained from the 29 interview participants to preserve their anonymity. At each participating healthcare facility, the consent process was witnessed by the head nurse, who did not participate in the interviews. Through the District Health Offices (DHOs), the NMEC informed all relevant healthcare facilities about this study before data collection.

### Data Analysis

Patient names and, where available, automatically generated case identification numbers were entered into a Microsoft Excel Spreadsheet for data management. After matching the paper form and the DHIS2 variables, the data were anonymized by removing the patients’ names. Analysis was done using the R programming language version 4.2.2 (19). The study used descriptive statistics to describe the level of completeness and timeliness of the MCIR 1-3-7 data collection forms. The study defined completeness (process) as the proportion of cases that had case- and foci investigations done. A case investigation and reactive case detection were deemed timely if executed within 3 days of the diagnosis, while foci investigation was considered timely when done within 7 days. Completeness of data was the proportion of forms filled with no blank spaces. The study did not assess the data completeness of the entomological investigation forms.

Voice recordings were transcribed verbatim and then translated into English. A codebook was developed with thematic codes based on the research question. The iterative process allowed the codes to be assigned to the data. Two coders independently reviewed all transcripts and assigned the codes. The codes were compared to identify similarities and differences in the interpretations. Reconciliation of the differences was also done to enhance the validity and reliability of the results and reduce bias. Quantitative and qualitative data were analyzed separately and integrated to assess how they complemented and confirmed each other. This approach provided a comprehensive understanding of the MCIR 1-3-7 from both a contextual and numerical perspectives.

## Results

### Quantitative assessment

A total of 510 malaria cases were retrospectively assessed in this study between 10^th^ and 20^th^ July, 2023, comprising 283 cases from Chikankata District and 227 cases from Mazabuka District. The monthly average case burden per facility was 28.3, with 95% CI [24.6, 32.0] in Chikankata and 25.2, 95% CI [16.9, 33.5] in Mazabuka.

The cases were classified according to the patients’ reported travel history. The MCIR 1-3-7 staff classified a total of 278 cases (54.5%) to have been imported from outside the district, province, or country, while 45.5% (232/510) of the cases were classified to have been transmitted locally. Forty-one (41) cases were not classified according to their place of origin, as the healthcare workers had not documented the travel history or preliminary classification of the cases’ origins. For this study, these cases were considered local. In addition, cases labeled as “local”, and “imported within district”, were also treated as local. Local cases (232, 45.5%) were eligible for foci investigation (Fig 4).

**Fig 4.**
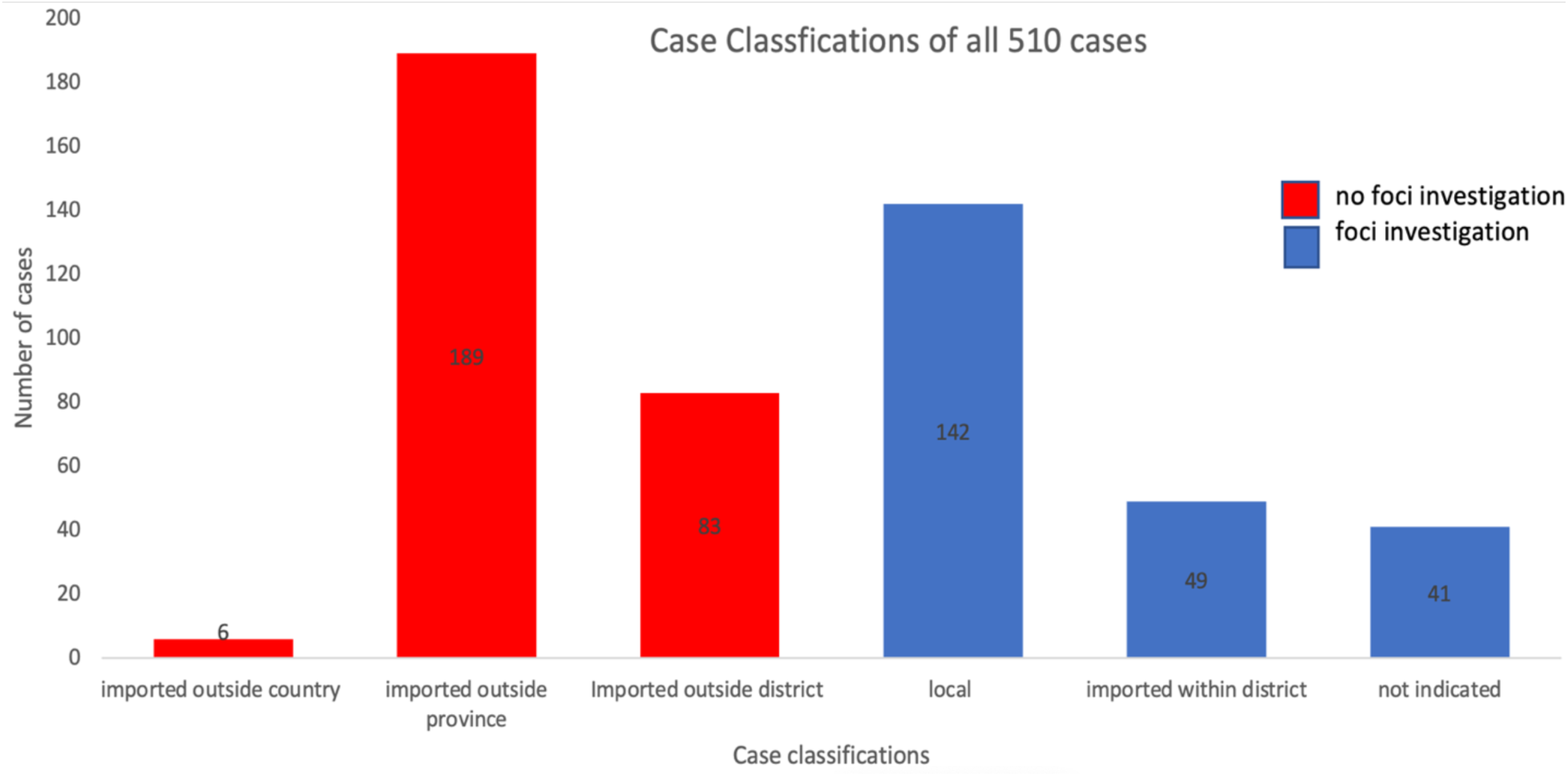
Case classification of the 510 sampled malaria cases.

Twenty-nine healthcare workers were interviewed, including 20 community health workers, seven environmental health technicians, and two nurses. Among the participants, 17 were males, and 12 were females. Three themes that included facilitators, barriers, and adoption of the MCIR 1-3-7 intervention were explored. These were based on the following intervention components: (i) case investigations and reactive case detection, (ii) foci investigations, (iii) form filling and data recording, and (iv) overall perception of the intervention implementation.

#### Case investigations and reactive case detection

A total of 139 case investigation form I were available at the sampled facilities. This represented 27.3% (139/510) reporting rate of these forms. This study excluded 12 forms due to issues such as incorrect or missing dates and a lack of identifiers. As a result, 127 forms were included in the analysis of timeliness and process completeness (Table 1), resulting in a 24.9% completeness rate for the 1-3 steps (127/510) among all malaria cases included in the study.

**Table 1.** Summary of assessed forms.

| Type of form | Case notification | Case investigation form I | Case investigation form II | Entomological investigation |
| --- | --- | --- | --- | --- |
| Total Number of forms found in facilities | 510 | 139 | 19 | 118 |
| Total number of forms expected | 510 | 510 | – | 232 |
| Number utilized in process completeness analysis | 510 | 127 | – | 90 |
| Number utilized in timeliness analysis | 510 | 127 | 15 | 86 |
| Number utilized in data filling completeness | 510 | 139 | 19 | – |

A summary of timeliness for the different activities is presented in Table 2. The study identified 19 case investigation form II, which were used to record reactive case detection data. Due to a gap in data collection, the study was unable to establish the number of reactive case detections that should have been conducted.

**Table 2.**
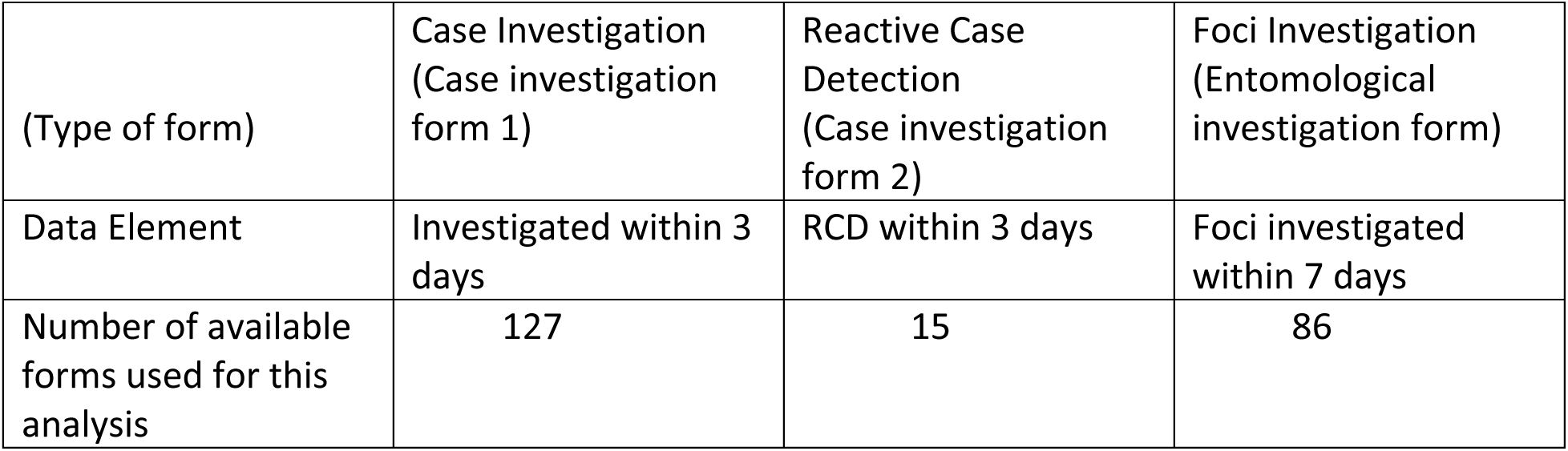
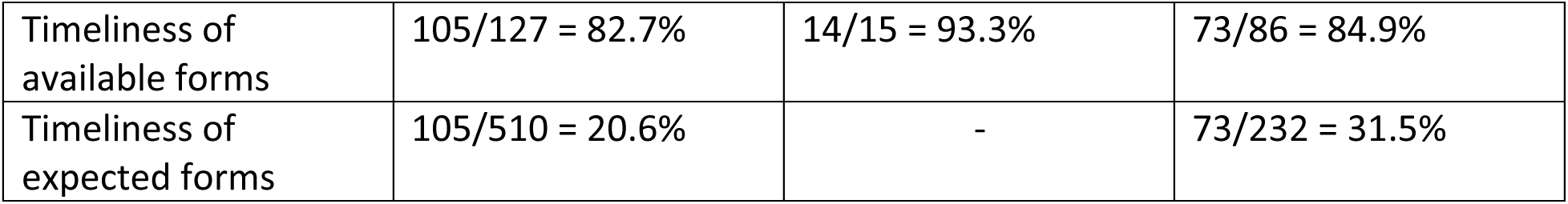
Timeliness of case and foci investigations.

| (Type of form) | Case Investigation (Case investigation form 1) | Reactive Case Detection (Case investigation form 2) | Foci Investigation (Entomological investigation form) |
| --- | --- | --- | --- |
| Data Element | Investigated within 3 days | RCD within 3 days | Foci investigated within 7 days |
| Number of available forms used for this analysis | 127 | 15 | 86 |
| Timeliness of available forms | 105/127 = 82.7% | 14/15 = 93.3% | 73/86 = 84.9% |
| Timeliness of expected forms | 105/510 = 20.6% | - | 73/232 = 31.5% |

Two factors facilitated the healthcare workers in achieving case investigations and reactive case detections. Collaboration within the team and coordinated patient visits, improved the efficiency of the healthcare workers.

> “We divide the work because day 3 has a lot of work…….so we help each other so that things happen quickly”. [Female CHW, Chikankata District]

The healthcare workers also facilitated their work by informing malaria patients about the MCIR 1-3-7 process and the subsequent visits they planned to make to the index case’s home. They further made appointments with index cases.

> “We make appointments with them prior to going…. So, most of the time, you (sic) usually find them”. [Female EHT, Chikankata District]

Several factors impeded the healthcare workers from conducting case investigations and reactive case detections. The healthcare workers complained about the distance they had to travel to implement this activity. They also tied this to a lack of transportation and protective clothing. Community health workers had to travel long distances on foot to reach the index case’s home. A male community healthcare worker in Chikankata District revealed that:

> “Challenges; we don’t have bicycles. We don’t have overalls.”

Competing interests also strained case investigation activity. The peak malaria season coincides with periods of abundant rainfall, during which families often engage in arable farming activities.

> “So, you find that the majority of the people are in the fields…they go and stay in the bush until time of (planted crop) harvest, so it’s also a challenge”. [Male CHW, Chikankata District]
>
> “If they are not in the fields, they’ve gone to fetch firewood. They are busy with other businesses. You know, charcoal making.” [Female EHT, Mazabuka District]

#### Foci investigations

One hundred and eighteen (118) Entomological Forms were found in the healthcare facilities, leading to a reporting rate of 50.9% (118/232). Only 90 forms were used to determine whether the foci-investigation step (-7 step) was done. The forms that were excluded had missing case classifications or investigation dates, as well as delays in foci investigations. This indicates that foci investigations were conducted for 38.8% (90/232) of eligible cases. To determine the timeliness of the foci investigation step, only 86 forms were utilized. Excluded forms had incorrect dates, missing case classifications or investigation dates, and delays in foci investigations. Only 31.5% (73/232) of the foci were investigated timely.

No facilitating factors were identified for conducting the foci investigations. The healthcare workers cited challenges, mainly long travel distances and a lack of transportation options. A male community health worker in one of the healthcare facilities, referring to his female colleague, pointed out that their health was put at risk for conducting this activity:

> “Yeah, even when she was pregnant, we used to walk almost 20km.”

In some healthcare facilities where motorbikes were available, foci investigations were hindered by insufficient fuel supplies, limiting the utilization of the bikes.

> “Sometimes, you find I have challenges with no fuel. I have the motorbike, but I cannot move.”
>
> [Male EHT, Chikankata District]

#### Form filling and data recording

Only 151 of the 510 (29.6%) case notification forms were filled out completely with all required components of the form completed, while 80 of the 139 (57.6%) case investigation form I were filled out completely. Out of the 19 case-investigation form II assessed, 36.8% (7/19) were completed in full. In this study, it was not possible to validate whether the data entered were correct.

The healthcare workers pointed out that they occasionally did not have paper data entry forms, in some instances for more than six months, nor could they afford to photocopy forms themselves. The healthcare workers documented the data on paper forms and subsequently entered the same data into the DHIS2 electronic surveillance platform. A review of this data revealed that 71.4% (364/510) of the case notification paper forms could be matched to data on the DHIS2 platform. These cases were further reviewed to ensure the contents matched precisely with the data entered on the paper forms. Of the 364 cases found on the DHIS2, only 16.8% (61 of 364) matched exactly with the data written on the paper forms. Most discrepancies appeared in case classification (19.2%), differences in patient names (12.4%), and the date of diagnosis (11.5%).

Some facilities used cellular phones instead of tablets to enter patients’ data into DHIS2. This arrangement had multiple concerns, ranging from non-functional devices to insufficient airtime to cover the data package. In Mazabuka District, a male community health worker lamented that:

> “No (cellphone) data. They don’t send us any”.

Healthcare workers could not transmit malaria patients’ data because they either did not have access to functional devices, or cellphone data bundles to access the electronic DHIS2.

#### Implementation of the MCIR 1-3-7 intervention

Multiple factors emerged as barriers to implementing the MCIR 1-3-7 intervention, with participants highlighting key challenges encountered during execution. They also made suggestions that could contribute to the success of MCIR 1-3-7 as an intervention.

In all the facilities, the interviewees mentioned that a lack of transport was at the top of the list as a challenge. This had indeed been cited as a problem when conducting both case- and foci investigations.

> “Transport, I think that’s one of the main issues. For me, it has been transport.”
>
> [Male EHT, Chikankata District]

In addition, the healthcare workers had proposed solutions to address this challenge.

> “What can make our job easy is transport, because even long distances will feel shorter because we have something to move with.” [Male CHW, Chikankata District]

A poor supply chain was also noted as a concern. There was a shortage of Rapid Diagnostic Tests (RDT), Personal Protective Equipment (PPE), Indoor Residual Spraying (IRS) insecticide, larvicide, and bed nets. The healthcare workers indicated that these shortages negatively affected the implementation of the MCIR 1-3-7 intervention.

> “At the moment, we don’t have the (bed) nets for MCI. So, we end up using the ones for MIP….”
>
> [Female EHT, Chikankata District]

Other healthcare facilities had a challenge with larvicide stock-out.

> “Chemical for larviciding was not there… and you scoop larvae, and then what is on your mind is, I need to do larviciding (sic). But how am I going to do it? There’s no chemical.”
>
> [Male EHT, Chikankata District]

The healthcare workers were of the view that the NMEC should provide all malaria supplies to ensure the successful implementation of the MCIR 1-3-7 intervention.

The healthcare workers reported low morale and a lack of motivation, particularly among community healthcare workers, who were performing volunteer work with no incentives attached. A male community healthcare worker in one of the facilities in Chikankata District lamented that:

> “We are volunteering. It’s hard work…. Then you are there…. You are walking for these long distances, but I am just volunteering for free, but we still do the work (sic).”

To overcome this challenge, the healthcare workers suggested that remuneration should be introduced as an incentive for MCIR 1-3-7 work. Even other healthcare workers who were salaried felt they could do more if there were incentives attached to the MCIR 1-3-7 work.

> “Motivation stipends for the CHWs ….. Even for the staff, the lunch allowances for every case, maybe when they go out there, so that they eat something.” [Male EHT, Chikankata District]

The healthcare workers further alluded that mentoring and supervisory visits to provide support, and problem-solving opportunities would help in implementing the MCIR 1-3-7 intervention. A healthcare worker pointed out that:

> “I think frequent monitoring and supervision, both from the district level, and the province level. Then maybe as well, the national level should be harnessed so that the program is effectively done.” [Male EHT, Chikankata District]

## Discussion

This is the first study to assess the MCIR 1-3-7 pilot project in Zambia. The study assessed the completeness, and timeliness of a sample of malaria cases in two low-incidence districts in the Southern Province between the 10^th^ and 20^th^ July, 2023. A total of 510 cases were retrospectively assessed in the 19 healthcare facilities visited. Additionally, the study assessed the perceptions of 29 healthcare workers across 11 healthcare facilities regarding enabling factors and hindrances to implementing MCIR 1-3-7 effectively. This study captured unfiltered responses that provided the first insights into the experiences of healthcare workers in implementing the MCIR 1-3-7 pilot project in Zambia. This area had never been explored before.

Most malaria cases registered in the two districts originated from outside the districts, as reported by the patients. Consequently, the healthcare workers classified the malaria cases as imported, assuming that the patients acquired the infections during travel. This finding aligns with surveillance data suggesting low local malaria transmission in the province, consistent with the classification of both districts as approaching elimination. The WHO Malaria Surveillance, Monitoring & Evaluation Reference Manual (20), and the Second Edition (21) highlight this trend, noting that as the proportion of local cases declines, the proportion of imported cases tends to increase, accompanied by a decrease in the total number of cases. *Kandel and colleagues*(2024) (22), reported that in Nepal, the number of indigenous cases decreased over the years, indicating epidemiological control of malaria transmission. This was also accompanied by a 40-58% increase in imported cases, while the overall total number of malaria cases declined.

Across all the interviews, the participants seemed eager to execute the activities of the surveillance intervention. However, challenges such as a lack of transportation and monetary incentives presented significant obstacles. Although the barrier of transportation has been noted before by *Silumbe and colleagues*(2015) (23) in their study on mass testing and treatment campaigns conducted by community health workers in Southern Province, the NMEC has the opportunity to strengthen the MCIR 1-3-7 intervention through the provision of bicycles. This measure would enhance the efficiency and timeliness of both case and foci investigations, thereby improving the overall surveillance intervention.

Supportive supervisory visits have been proven to improve ‘worker performance’ (24) as well as data quality that includes collection, reporting, and documenting (25). According to *Bello and colleagues*(2013) (24) “supervision has been reported as one of the nonmonetary motivators of health workers and its absence can be correlated with health workers’ poor job satisfaction as well as poor service delivery”. Working through the district health offices, the NMEC should leverage this approach to boost implementer morale. Beyond improving data quality - particularly timeliness, completeness, form filling, and data recording - this strategy helps close existing knowledge gaps. It further enables monitoring of the MCIR 1-3-7 intervention while offering mentorship, clarification, and reinforcement of best practices to enhance healthcare workers’ confidence.

Refresher training courses have been found to significantly improve retention of skills (26),(27). Tetteh *and colleagues*(2021) (27) in their study assessing the impact of a supplementary training program among malaria laboratory personnel indicated that refresher trainings have a significant impact on the accuracy of malaria microscopy and achieves exceptional outcomes. While the NMEC’s support for MACEPA-led training of healthcare workers on MCIR 1-3-7 principles is commendable, sustained investment in refresher trainings is essential to maximize the benefits of this initial effort. This measure would ensure a strengthened quality of service delivery and enhance surveillance. In addition, further investments by the NMEC are needed to provide job aids (flip charts, posters, reference cards, flowcharts) that support healthcare workers in performing tasks more effectively and accurately.

To ensure successful and optimal implementation, the NMEC should standardize and clearly define key surveillance terms, including index case, local case, imported case, foci, and reactive case detection, to reduce ambiguity and confusion among implementing healthcare workers. Furthermore, to strengthen surveillance processes, the NMEC should provide clear guidance through standard operating procedures (SOPs) specifying that all cases require investigation and follow-up, the appropriate timing of such investigations, and the circumstances under which foci investigations should be conducted. Such clarification is also expected to improve reporting rate.

Given the positive step that the government of Zambia allocated 5.26% of the GDP to the Ministry of Health in 2022 (28), and the NMEC committed nearly US$2 million to the malaria budget in 2024 (6), significant funding gaps remain. This was evidenced by reports from healthcare workers indicating frequent stockouts of various malaria commodities. The End Malaria Council should therefore strengthen resource mobilization to enable the NMEC to procure malaria commodities (RDTs, drugs, PPE, LLINs, IRS insecticides, larvicides) and remunerate community healthcare workers. In addition, this funding will facilitate the continuation of malaria surveillance activities and data reporting mechanisms that had previously been supported by MACEPA until the end of 2023.

Although this study provides important insights, several limitations should be highlighted. The MCIR 1-3-7 was piloted in over 50 healthcare facilities in the two districts. However, the study visited only 19 healthcare facilities across the two districts. This resulted in a small sample size of the sampled cases. This study used a convenience sample, hence lacked generalizability to all 50 healthcare facilities. In addition, while qualitative research is not intended to produce generalizable findings, a greater sample size could have revealed a wider range of experiences relevant to the local context and identified diverse facilitating factors or barriers in implementing MCIR 1-3-7 activities.

This study could not ascertain whether malaria case notifications were done within 1 day. The researchers did not have access to cellphone short messages (SMS), emails, or telephone records to determine whether staff at the healthcare facilities sent notifications during the days corresponding to the dates of the patient’s visits. Furthermore, the researchers did not have access to the district health office’s malaria registers. Access to these registers would have helped verify the dates of case notifications for malaria patients, allowing comparison between the date of diagnosis and the report to the district health offices.

## Conclusion

While the MCIR 1-3-7 pilot project has faced implementation challenges, it presents significant opportunities for strengthening the intervention. Enhancing support supervisory visits, conducting regular data audits, and providing desk and wall job aids are practical steps NMEC can leverage. These measures would help address current gaps in timeliness, completeness, and data quality.

## Data Availability

De-identified data are available from the author on reasonable request.

## Declarations

### Ethics approval and consent to participate

Ethical approval was obtained from the University of Zambia Biomedical Research Ethics Committee (UNZABREC) and the National Health Research Authority (NHRA). An approval was granted on June 27, 2023 (REF. NO. 4100-2023).

### Consent for publication

Not applicable

### Authors’ contributions

Designed the study: TPE, RYK. Acquisition of the data: RYK, SMK. Analyzed the primary data presented in this paper: RYK, SMK, JAS, RAA, TPE. Wrote the first draft: RYK. All other authors reviewed and approved the final document.

## Acknowledgments

This study was made possible by a committed team of staff, data collectors, and community health workers. We also wish to thank the participating communities, health facilities, and district health teams for providing their support. We would like to extend our gratitude to the PATH Partners Group and the NMEC Zambia for their role in the partnership.

## Supporting information

### Supporting information 1

#### Interview guide

The researchers collected data through 11 questions designed explicitly for this study. The questions were open-ended and semi-structured to extract information related to the interviewees’ work, experiences, and views. The interviewees were asked the following questions:

- When did this health facility start implementing malaria case investigations using the 1-3-7 approach?
- What type of training did you all receive in the malaria case investigations using the 1-3-7 approach?
- Can you all please describe to me what malaria case investigations using the 1-3-7 approach entail?
- Are there any challenges with completing the Case Notification Form within 1 day of the case being entered into the CHW or health facility log-book? If so, what are some of the challenges?
- Are there any challenges with completing the case investigation within 3 days of identification of the index case, and completing the Malaria Case Investigation Form I? If so, what are some of the challenges?
- Are there any challenges with testing and identifying secondary cases around the index case, and completing the Malaria Case Investigation Form II? If so, what are some of the challenges?
- What are the challenges with completing the Entomological Investigation Form within 7 days from an identified index case? *[Can remind them this is supposed to be completed by the environmental health technician and is used to collect information on the foci within a 2-kilometer radius of the index case, including the number and location of potential larval sources.]*
- Are there any challenges in implementing responses in a focus, such as distributing insecticide-treated bednets or performing indoor residual spraying? If so, what are the challenges? ***[Prob]*** Are there ever any stock-outs?
- Has the health facility started performing any larval source management, environmental management, or fish that eat mosquito larvae? If so, what have been the primary challenges with using these responses? If so, how have the communities reacted to these types of responses?
- Do you all do any sort of communication (or social behavior change communication) in the community around an index case? If so, what does this entail? Are there any issues with doing this malaria communication?
- If you could name three things that would help you implement malaria case investigations using the 1-3-7 approach, what would they be?

## Supporting information 2

### Detailed description of how the MCIR 1-3-7 system is supposed to be implemented

#### Day 1

Healthcare workers are supposed to test all individuals who present at the healthcare facilities with suspected malaria. Confirmed cases are reported to the next higher level in the Ministry of Health hierarchy within 24 hours. If a case is diagnosed in the community by a community healthcare worker, it must be reported to the healthcare facility and to the District Health Office. A case diagnosed at the healthcare facility is reported to the District Health Office (DHO). The healthcare workers report the case to the higher level through a cellphone text message or a voice telephone call. The patient is supposed to be treated with Artemisinin-based Combination Therapy (ACT) or referred (for further management) according to the severity of their illness. A low-dose primaquine is administered to patients who have been prescribed oral medication. The preliminary classification of the case as locally acquired or imported is done based on clinical presentation, past medical history, and the patient’s recent travel history. A dried blood spot (DBS) should be collected for molecular surveillance, enabling the genotypic characterization of anti-malarial drug-resistance markers, and parasite strain identification.

Documentation of the case is entered on the paper case notification form and the DHIS2 platform. Each patient’s unique ID case number is generated in DHIS2 and should be manually copied to the paper case notification form. Plans for the Day 3 home visit are communicated to the patient.

#### Day 3

The MCIR 1-3-7 staff are supposed to visit the index case’s home during the case follow-up process. The healthcare workers review the patient’s health status, inspect medication packets to assess adherence to the prescribed treatment, and verify whether a DBS has been collected. During the visit, further inquiry is made into the patient’s travel history, and the possibility of importation of the infection is explored. During this visit, the healthcare workers are supposed to make a definitive case classification. Members of the family are tested for malaria with RDTs. Details are entered into the case investigation form I and DHIS2. The healthcare workers also conduct RDT testing for neighbors within a 140-meter radius. Individuals who test positive for malaria must be treated according to the treatment protocol (with ACT and primaquine or referred). Details of each new case are recorded in a case notification form. These cases must be reported to the higher level. A unique ID case number is generated for each new patient. Each patient’s details are recorded on the DHIS2 platform. Plans are also made to conduct case follow-up for these patients after three days. Cases verified to have been contracted within the district are reported to the healthcare facility nearest the suspected transmission location. A system of referral and collaboration between healthcare facilities is utilized for this process.

During this visit, the healthcare workers are supposed to assess the home’s surrounding environment to identify factors that support malaria transmission. These include checking for potential mosquito breeding sites such as standing water, improper water storage practices, and drainage. They also assess household structures for open eaves and the presence or absence of window screens, as well as environmental management practices that include vegetation around the home and garbage disposal. The team also starts plans to curb the transmission.

#### Day 7

For cases confirmed to be locally transmitted, the MCIR 1-3-7 staff are supposed to conduct another follow-up visit to conduct an entomological investigation and implement a response. The response measures may include reactive targeted Indoor Residual Spraying (IRS), distribution of Long-Lasting Insecticidal Nets (LLINs), environmental management activities such as vegetation clearance, burying or larviciding of breeding sites, dissemination of health promotion messages, and implementation of structural interventions like screening on windows and eaves to reduce vector entry. Information on breeding sites and the presence of mosquitoes is collected. The staff also records GPS information of the locale and classifies the focus accordingly (active, residual non-active, or cleared). In addition, the staff collects data on the coverage of vector control measures within a 2-kilometer radius. These include data on the number of structures eligible for IRS, the number of structures covered with IRS in the preceding 12 months, the quantity of LLINs distributed in the targeted area over the same period, and the proportion of individuals reporting to have slept under an LLIN the previous night. Data is entered on the paper entomological investigation form and the DHIS2.

## Supporting information 3

### Case Notification Form

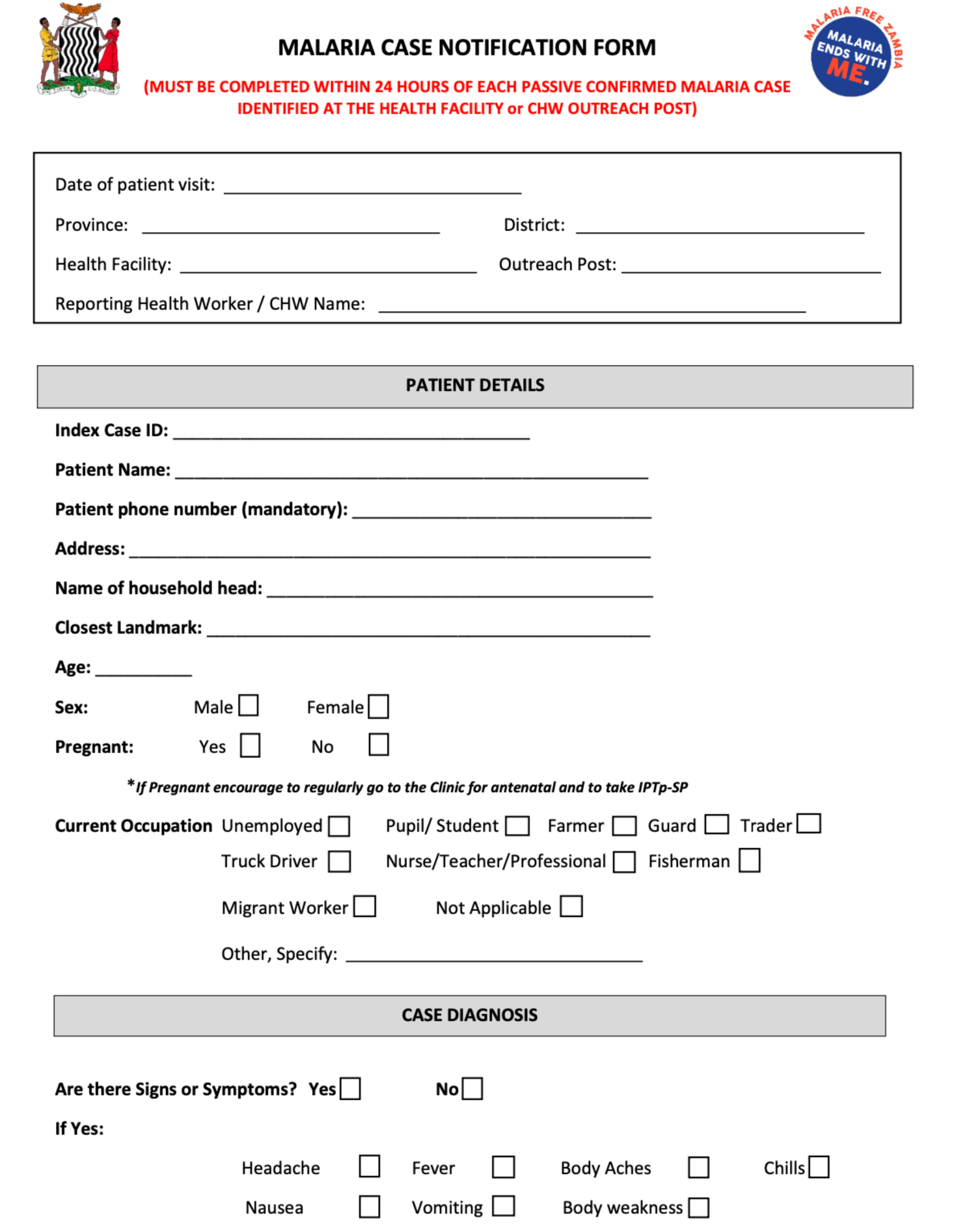
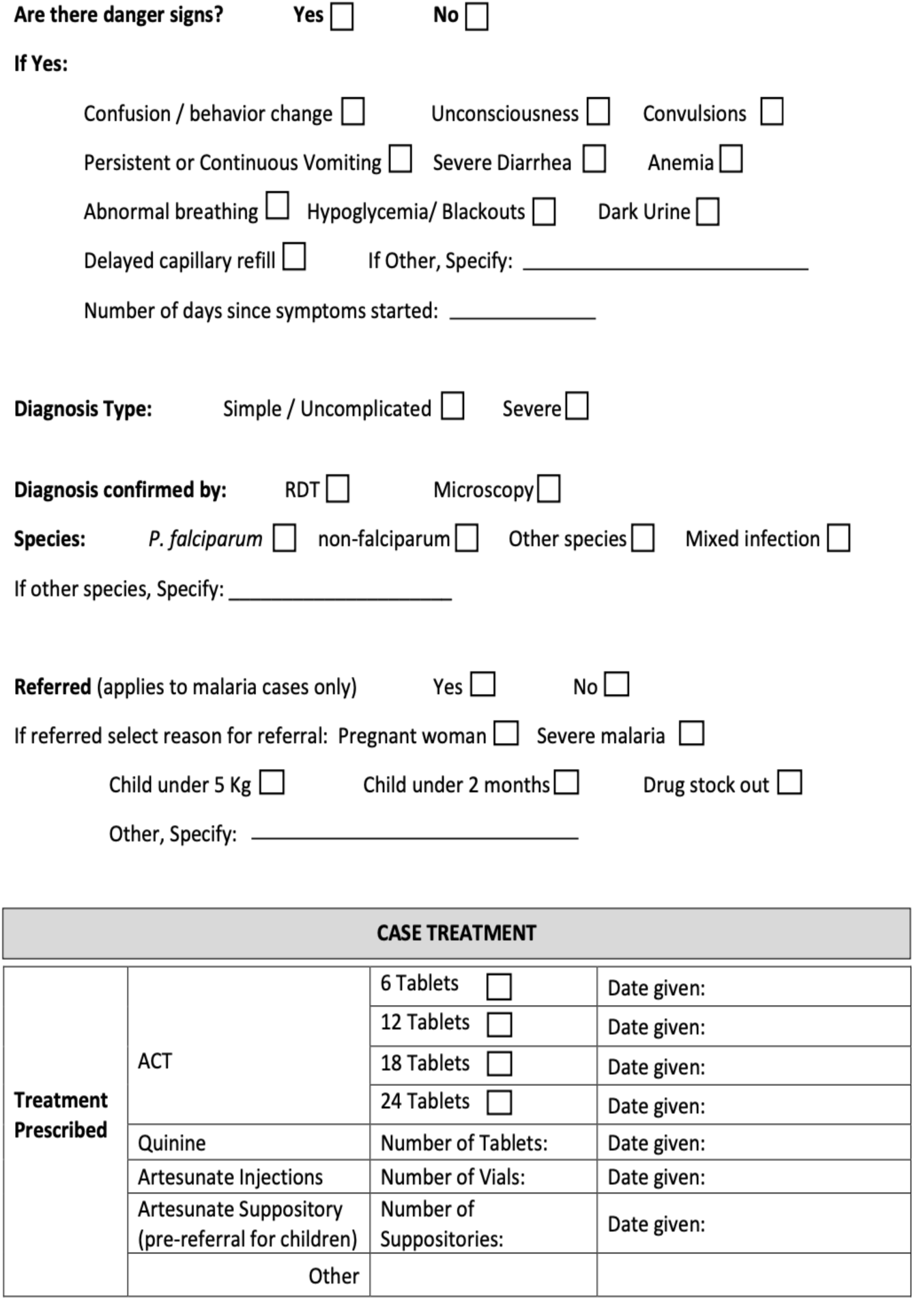
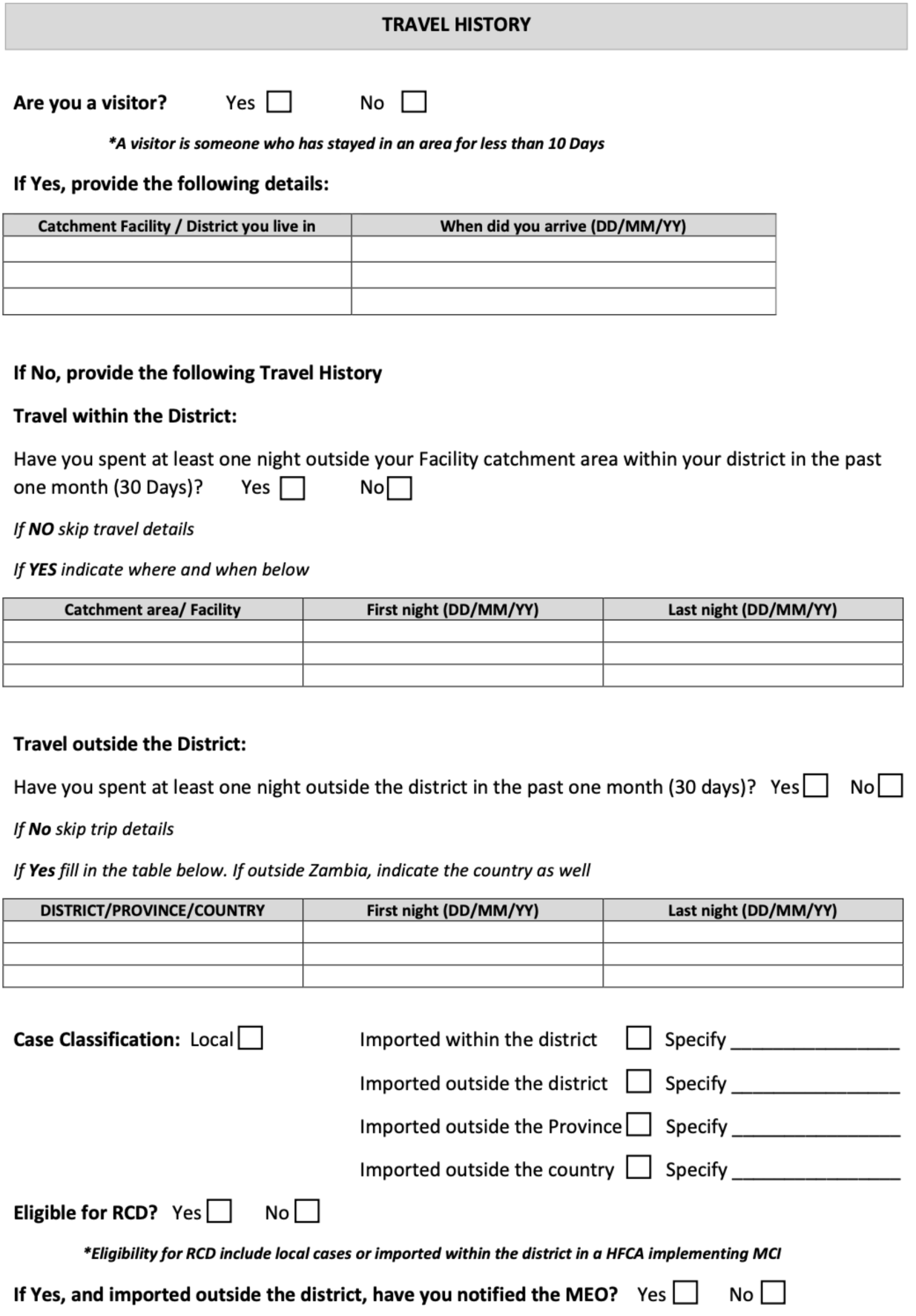
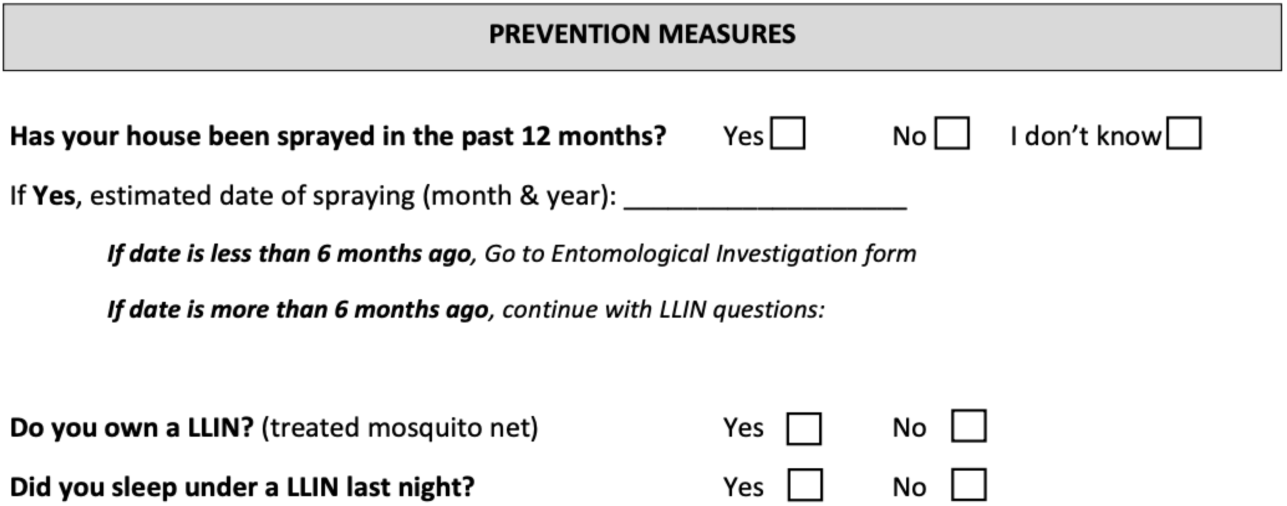

## Supporting information 4

### Case Investigation Form 1

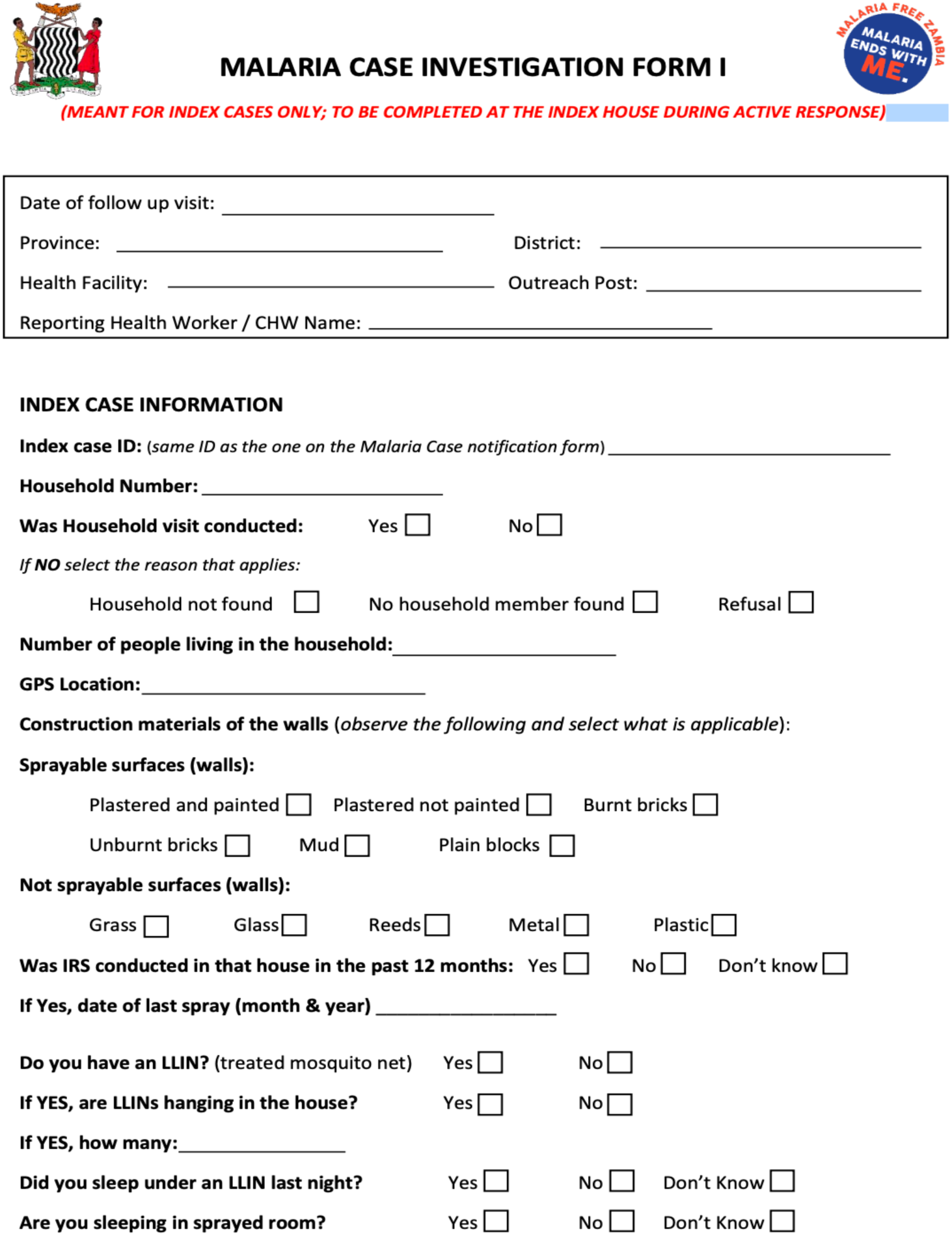
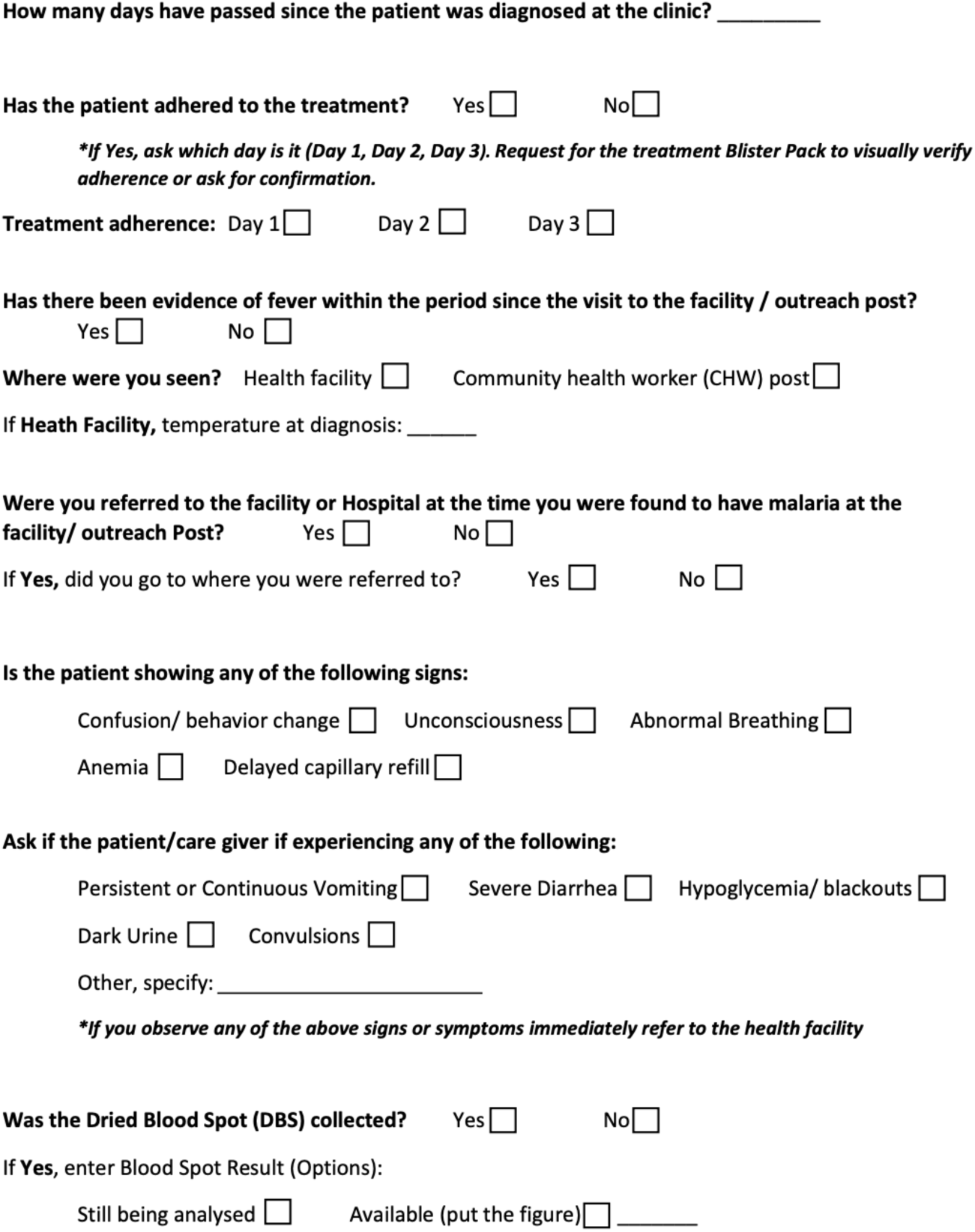

## Supporting information 5

### Case Investigation Form 2

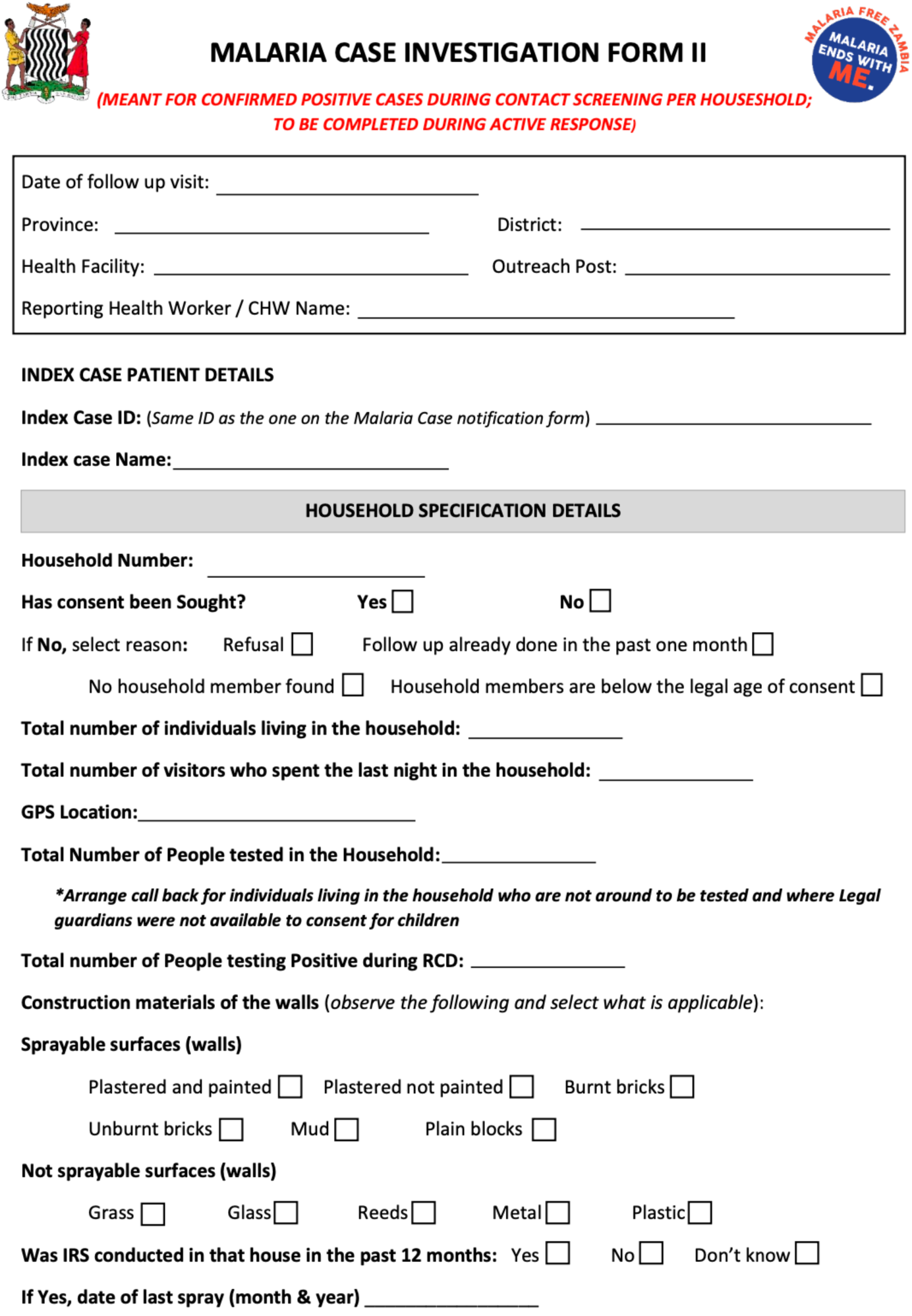
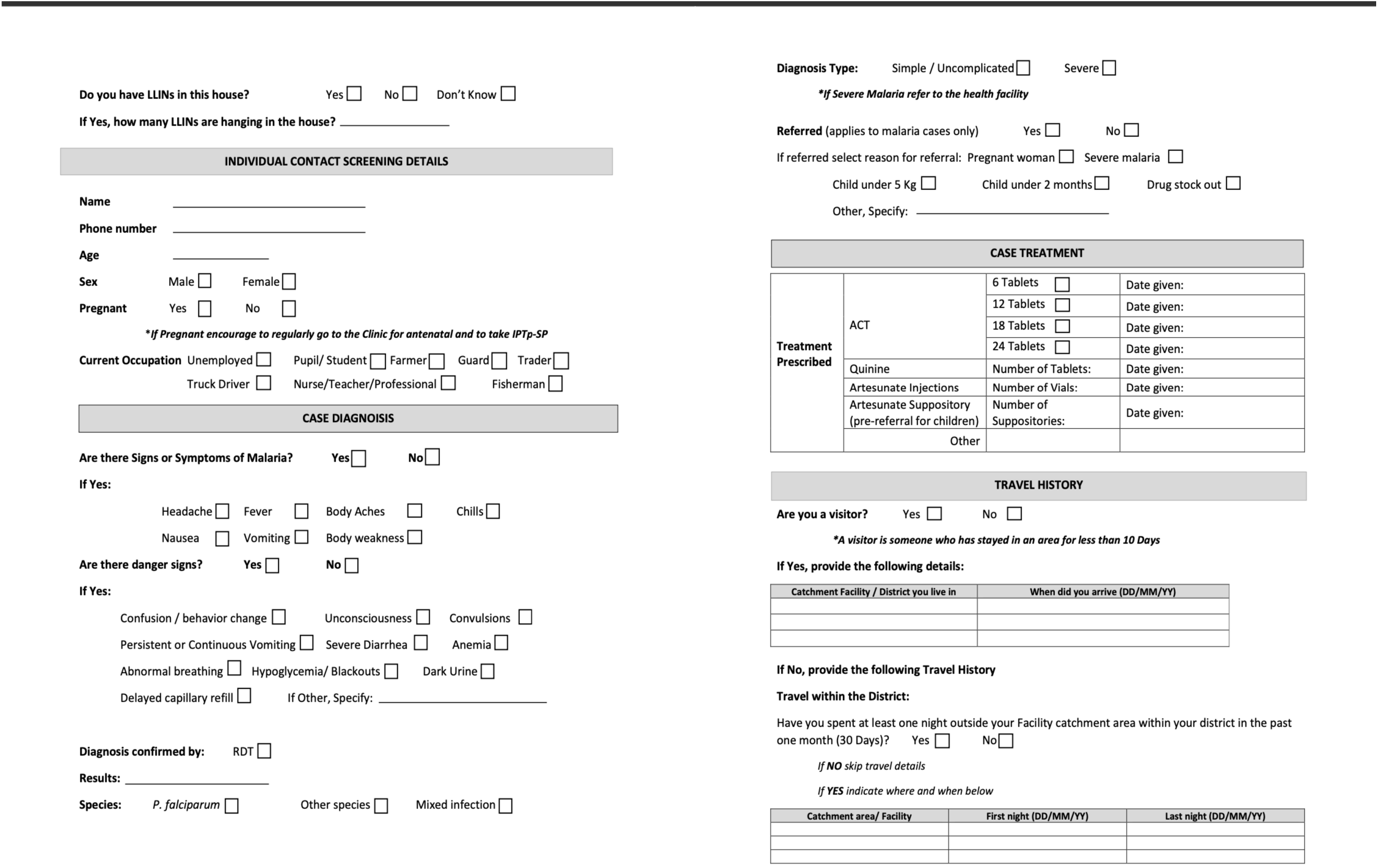
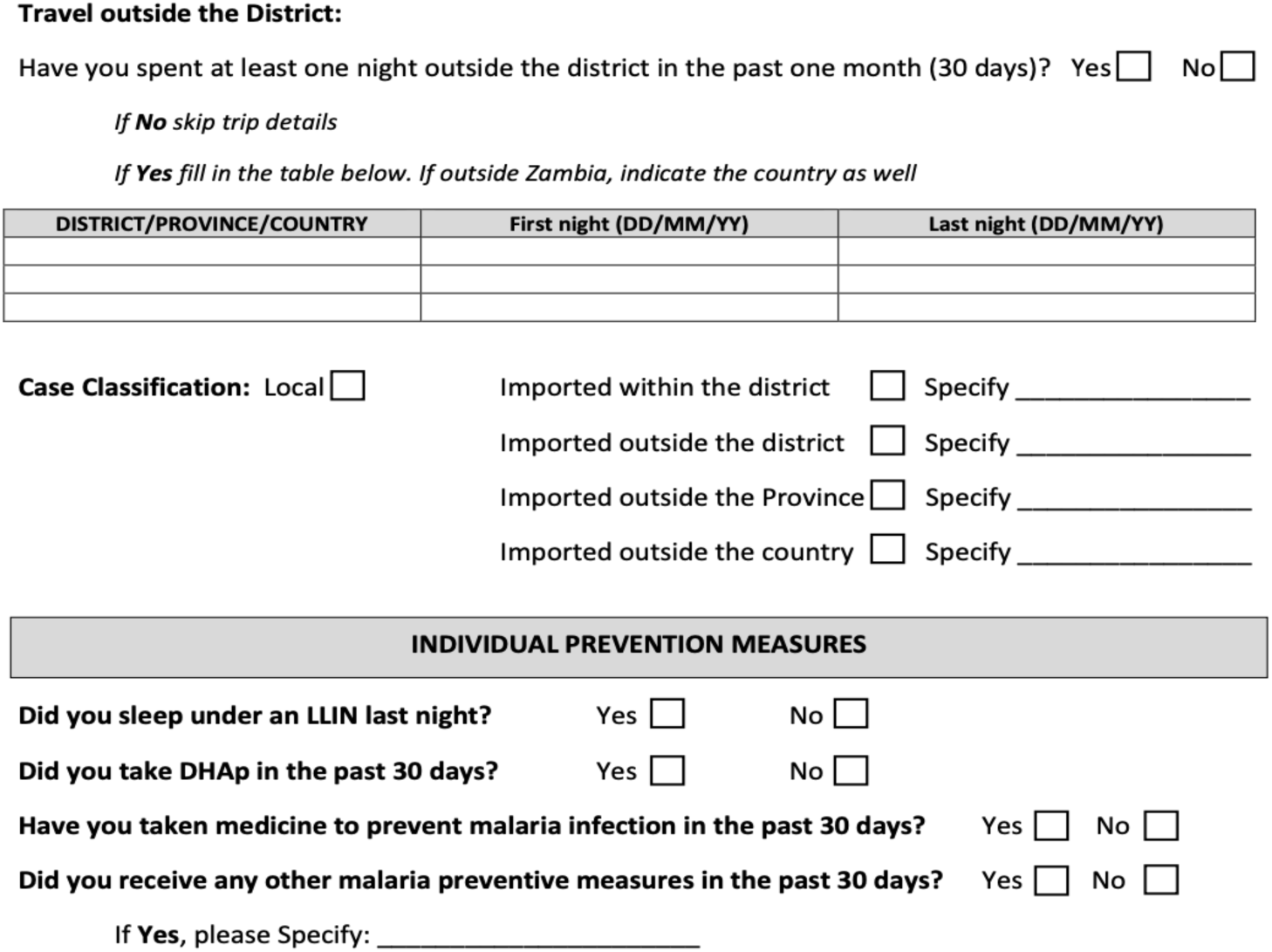

## Supporting information 6

### Entomological Investigation Form

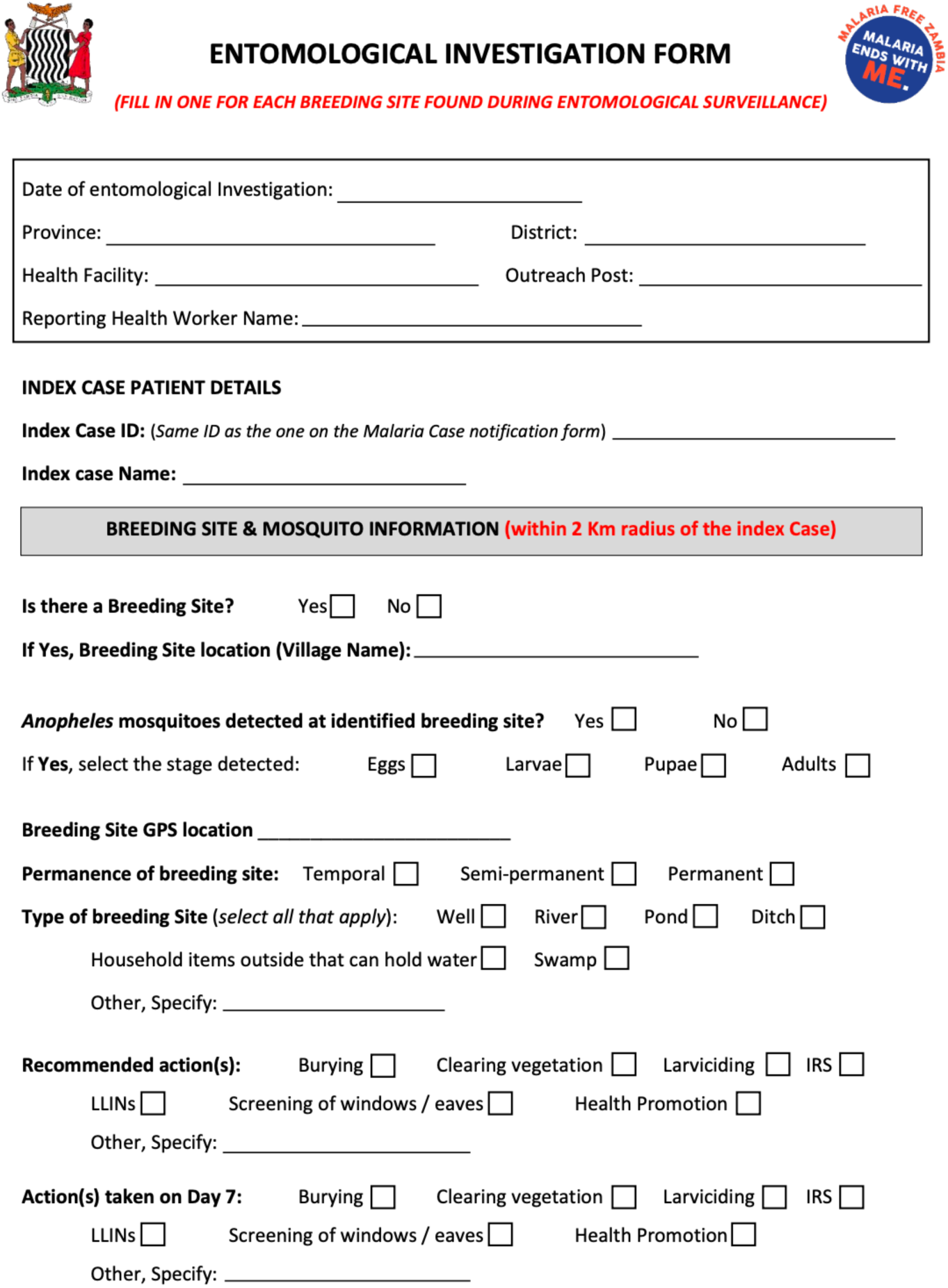
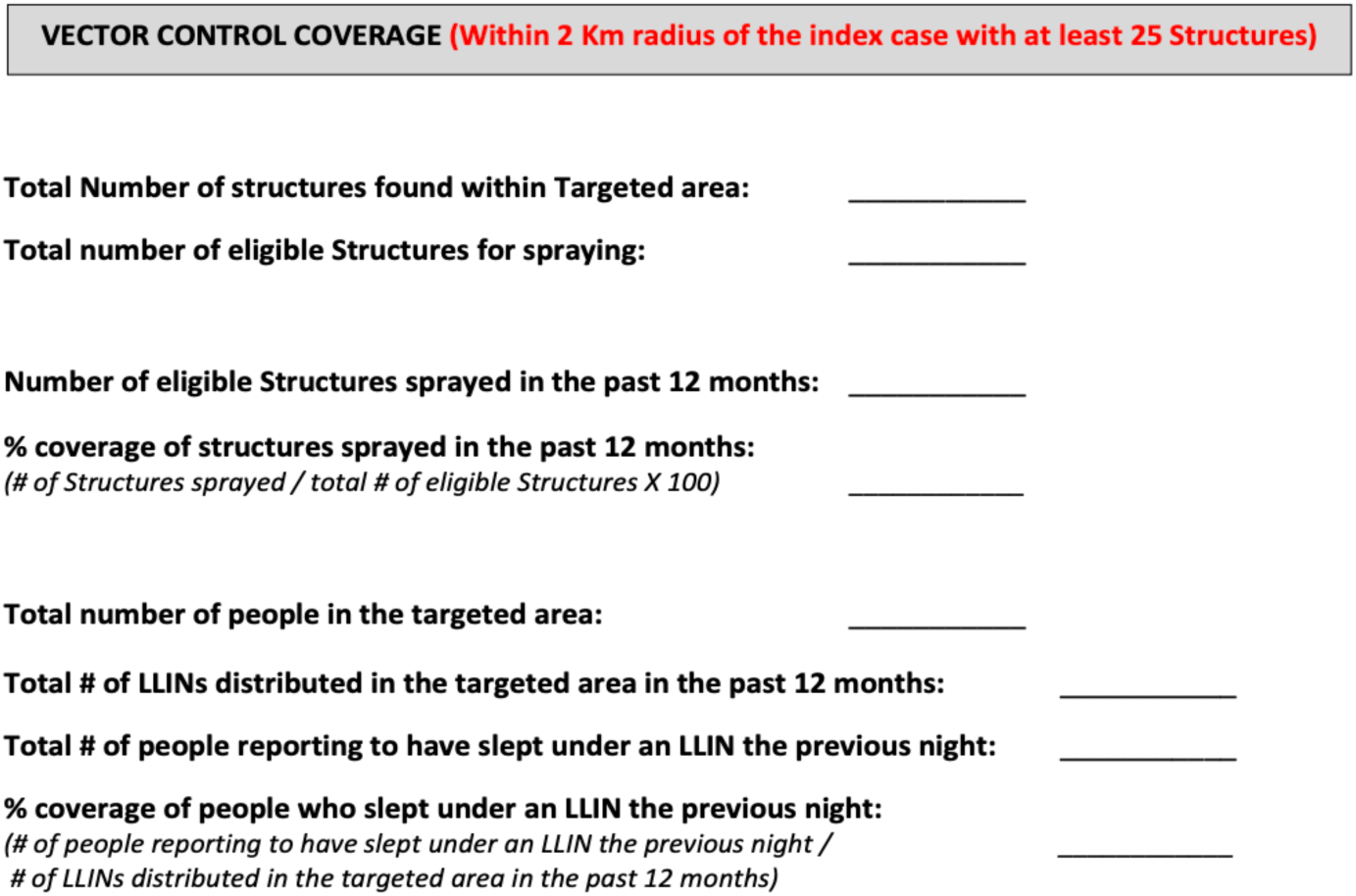

